# Reducing the Burden of Hypoglycemia: FLO23011 Improves Patient-Reported Outcomes and Identifies Glycemic Predictors of Treatment Success

**DOI:** 10.64898/2026.07.29.26359225

**Authors:** David Russell-Jones, Eliza G. Meehan, Vera Smout, Sabyasachi Roy, Wendy Frost, Tim M. Young, David B. Bartlett

## Abstract

**Objective:** To compare patient-reported outcomes (PROs) for hypoglycemia recovery with FLO23011, a glucose/beta-hydroxybutyrate Multi-Energy Substrate for Hypoglycemia (MESH) treatment, versus standard glucose gel in type 1 diabetes, contextualized by continuous glucose monitoring (CGM) analyses.

**Research Design and Methods:** In a randomized, open-label, crossover study, 12 adults with type 1 diabetes used FLO23011 or glucose gel to treat hypoglycemia during two 6-week CGM-monitored periods. PROs were assessed using 14-domain questionnaires and exit interviews. CGM analyses from a broader discovery analysis examined patient-relevant recovery signals: glucose-band exposure versus psychological PRO scores, and baseline glycemic variability versus time-in-range response.

**Results:** FLO23011 was rated more favorably than glucose gel in 13/14 domains, with 10 significant. Differences included speed of action (8.0 vs. 7.3; P = 0.025), after-effects reduction (8.0 vs. 6.2; P = 0.014), ease-of-use (8.9 vs. 4.9; P = 0.002), and overall management ability (8.5 vs. 7.4; P = 0.019). Interviews described faster cognitive recovery, reduced disruption, and greater confidence. Reduced Level 1 hypoglycemia exposure was associated with higher reduced-worry and management-ability ratings (ρ = 0.90; P = 0.037). Higher baseline coefficient of variation predicted greater time-in-range improvement with FLO23011 (ρ = 0.917; P = 0.0005).

**Conclusions:** FLO23011 showed more favorable patient-reported recovery than glucose gel. Initial CGM-PRO analyses findings suggest perceived benefit may align with reduced Level 1 hypoglycemia, while baseline variability may identify greater objective response. Results support integrating patient-reported and glycemic outcomes to evaluate hypoglycemia treatments, warranting confirmation in larger blinded studies.

## Introduction

Hypoglycemia remains a frequent and disruptive complication of insulin therapy in type 1 diabetes, with individuals experiencing up to 200 treatment-requiring episodes per year (1). Current guidelines recommend 15–20 g of fast-acting carbohydrate as first-line treatment (2). Although effective at restoring plasma glucose, glucose-only treatment may be followed by glycemic oscillations (3) and prolonged neuroglycopenic symptoms such as fatigue, impaired cognition, and reduced functional capacity (1,4–6). These sequelae disrupt daily life and contribute to fear of hypoglycemia and suboptimal disease management (7–11), yet treatment evaluation has focused predominantly on glycemic endpoints, with the patient experience of recovery remaining poorly characterized.

Convergent experimental evidence suggests that neuroglycopenic symptoms may persist after blood glucose has normalized because cerebral energy metabolism remains transiently impaired during recovery from hypoglycemia (12–15). One proposed mechanism is a transient hypoglycemia-related “energy bottleneck,” in which hypoglycemia-induced activation of poly(ADP-ribose) polymerase-1 (PARP-1) depletes cytosolic NAD⁺, constraining neuronal glycolytic flux (16–18). This energy bottleneck has been proposed as a potential contributor to the commonly reported “hypoglycemia hangover,” characterized by post-episode fatigue, cognitive fog, and functional impairment that can persist after blood glucose has returned to range (16).

FLO23011/ Klario (FLO) is a novel Multi-Energy Substrate for Hypoglycemia (MESH) recovery drink developed to address this recovery gap. It combines glucose with beta-hydroxybutyrate (BHB), an alternative substrate that can enter oxidative metabolism downstream of glycolysis, and may therefore support neuronal ATP production during the post-hypoglycemia window in which glycolytic flux may be constrained (16,19).

The pharmacokinetic outcomes, primary glycemic outcomes, and an initial summary of patient-reported outcomes from the FLO study have been previously reported (20). In that report, FLO was shown to significantly improve post-hypoglycemia time-in-range (TIR; +5.5 percentage points, P = 0.019) and reduce recurrent hypoglycemia within 2 hours by 27% (P = 0.031) relative to standard of care (SoC) glucose gel. Patient reported outcomes (PROs) were also summarized at a high level, with ratings favoring FLO in 13 of 14 assessed domains (20).

The present report provides the first integrated report of PROs from the randomized FLO study, including the full domain-level questionnaire dataset, direct product comparator ratings, and structured exit interview findings. To contextualize the patient-reported findings, we also report for the first time, focused continuous glucose monitoring (CGM) analyses derived from a broader discovery analysis of the study dataset, focusing on the signals most relevant to patient-centered hypoglycemia treatment, including associations between CGM-derived glucose band metrics and patient reported-outcomes, and between baseline glycemic variability and objective CGM-derived treatment response.

## Research Design and Methods

### Study design and participants

This was a randomized, open-label crossover study in 12 adults with type 1 diabetes (8 Male / 4 Female; mean age 49.3 ± 11.3 years; HbA1c 6.8% ± 0.8% [51.3 ± 8.9 mmol/mol]). Participants were enrolled between 12 November 2024 and 22 January 2025. The study design, participant eligibility criteria, CGM methodology, episode definitions, pharmacokinetic outcomes, primary glycemic outcomes, and an initial summary of PROs have been reported previously (20). The present analysis expands the patient-reported dataset from the same randomized trial and reports structured qualitative findings and CGM-PRO analyses.

Participants were randomized in a 1:1 ratio to use either FLO (15 g fast-acting carbohydrate + 10 g BHB) followed by SoC (glucose gel) containing 15 g glucose, or SoC glucose gel followed by FLO, to treat naturally occurring hypoglycemic episodes during normal daily life. Each treatment period lasted 6 weeks, with CGM throughout the full study period and no washout period. Participants continued their usual diabetes care during the study; concomitant care was not otherwise controlled. A total of 1,032 hypoglycemic episodes were identified from 232,607 CGM readings over 122 days of monitoring.

The study was approved by the Leicester Central Research Ethics Committee and was coordinated through CEDAR, Royal Surrey NHS Foundation Trust, Guildford, UK.

### Patient-reported outcomes

PROs were assessed using structured questionnaires across 14 domains evaluating clinical effectiveness, functional recovery, product characteristics, and psychological impact. At the end of each 6-week treatment period, participants rated the assigned product independently using a 10-point Likert scale, where higher scores indicated a more favorable rating for the product assessed.

At the end of the full 12-week study, direct comparator ratings were also obtained across the same 14 domains using a single anchored scale. Results are presented as the percentage of participants scoring above 5 (indicating preference for FLO).

Structured exit interviews were conducted at study completion to capture participants’ experiences of hypoglycemia treatment and recovery with each product. Closed-response items were summarized descriptively as percentages. Open-ended responses were reviewed using an inductive thematic approach. Recurring concepts related to treatment experience and hypoglycemia recovery were identified from participant responses and grouped into higher-order themes. Themes were reviewed and illustrated using representative quotations.

### Focused CGM analyses

CGM analyses were conducted after quality control and data harmonization. Focused analyses examining associations between CGM-derived metrics and patient-reported recovery outcomes were supported by KOSMOS (Edison Scientific), an AI-assisted analysis tool. KOSMOS generated CGM-PRO analysis outputs, which were manually reviewed and verified before inclusion in the final analyses. Participants were included in these analyses if paired CGM data were available from both treatment periods. Treatment-associated differences across CGM-derived glucose band metrics and psychological PRO domain scores were assessed. Glucose bands examined included the low-normal range (3.9–5.0 mmol/L) and the Level 1 hypoglycemia range (3.0–3.8 mmol/L). Assessment included whether baseline glycemic variability predicted objective glycemic response to FLO. Baseline variability was expressed as coefficient of variation during the SoC period, and treatment response was defined as the within-participant difference in CGM-derived TIR between FLO and SoC.

### Statistical analysis

PRO data were compared using linear mixed-effects models (statsmodels, Python), controlling for participant and period effects. P values were corrected using the Benjamini–Hochberg procedure; significance was defined as false discovery rate (FDR) <0.05. Cohen’s d was used to report effect sizes. CGM-PRO and baseline predictor analyses used Spearman rank correlation. Given the small sample size and hypothesis-generating nature of these analyses, no correction for multiple comparisons was applied.

## Results

### Full Domain-Level Patient-Reported Outcomes

The complete domain-level PRO analysis, expanding the previously summarized PRO findings, is shown in Table 1. Mean scores were higher for FLO than SoC in 13 of 14 domains, with 10 statistically significant after correction for multiple comparisons. Significant differences included speed of action (8.0 vs 7.3, P = 0.025), reduction in after-effects (8.0 vs 6.2, P = 0.014), and overall hypoglycemia management ability (8.5 vs 7.4, P = 0.019). Effect sizes were moderate to large across these domains. Ease of use during hypoglycemia showed the largest effect (8.9 vs 4.9, P = 0.002).

**Table 1.**
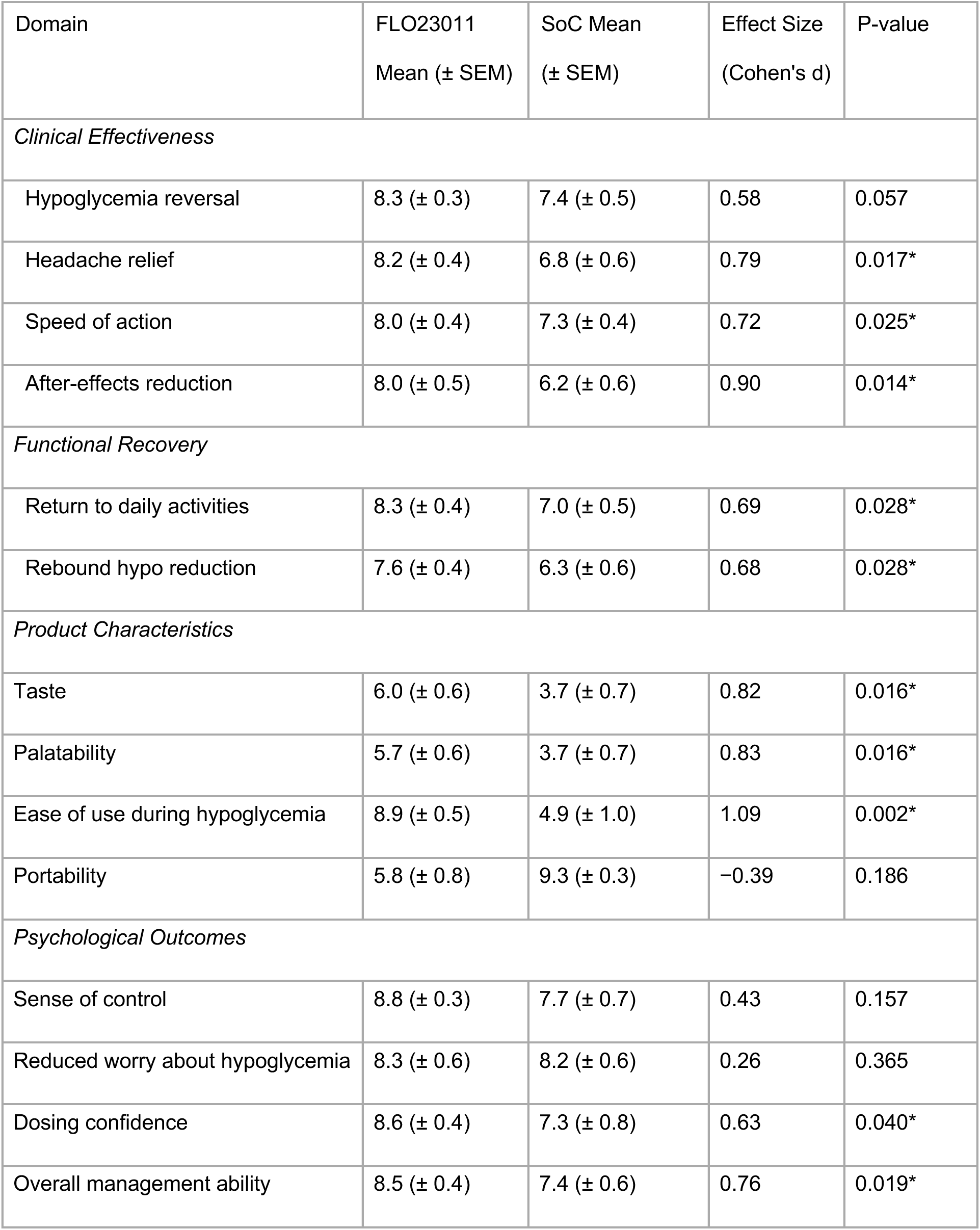
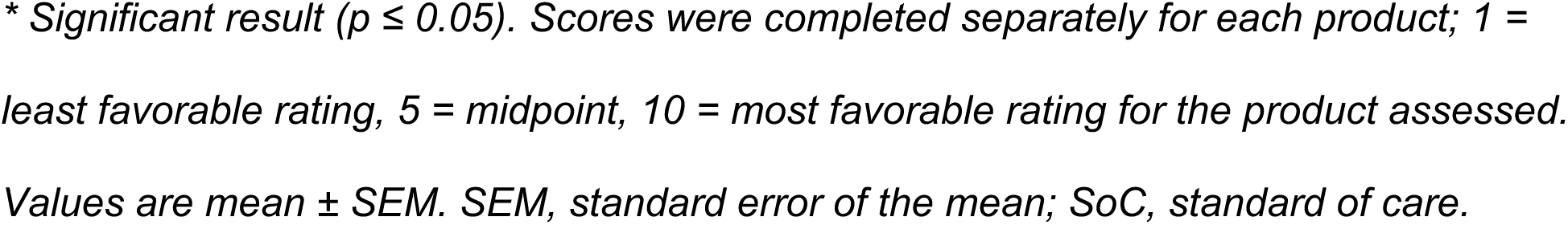
Full domain-level patient-reported outcome scores: FLO23011 vs Standard of Care.

| Domain | FLO23011<br>Mean (± SEM) | SoC Mean<br>(± SEM) | Effect Size<br>(Cohen's d) | P-value |
| --- | --- | --- | --- | --- |
| *Clinical Effectiveness* |  |  |  |  |
| Hypoglycemia reversal | 8.3 (± 0.3) | 7.4 (± 0.5) | 0.58 | 0.057 |
| Headache relief | 8.2 (± 0.4) | 6.8 (± 0.6) | 0.79 | 0.017* |
| Speed of action | 8.0 (± 0.4) | 7.3 (± 0.4) | 0.72 | 0.025* |
| After-effects reduction | 8.0 (± 0.5) | 6.2 (± 0.6) | 0.90 | 0.014* |
| *Functional Recovery* |  |  |  |  |
| Return to daily activities | 8.3 (± 0.4) | 7.0 (± 0.5) | 0.69 | 0.028* |
| Rebound hypo reduction | 7.6 (± 0.4) | 6.3 (± 0.6) | 0.68 | 0.028* |
| *Product Characteristics* |  |  |  |  |
| Taste | 6.0 (± 0.6) | 3.7 (± 0.7) | 0.82 | 0.016* |
| Palatability | 5.7 (± 0.6) | 3.7 (± 0.7) | 0.83 | 0.016* |
| Ease of use during hypoglycemia | 8.9 (± 0.5) | 4.9 (± 1.0) | 1.09 | 0.002* |
| Portability | 5.8 (± 0.8) | 9.3 (± 0.3) | -0.39 | 0.186 |
| *Psychological Outcomes* |  |  |  |  |
| Sense of control | 8.8 (± 0.3) | 7.7 (± 0.7) | 0.43 | 0.157 |
| Reduced worry about hypoglycemia | 8.3 (± 0.6) | 8.2 (± 0.6) | 0.26 | 0.365 |
| Dosing confidence | 8.6 (± 0.4) | 7.3 (± 0.8) | 0.63 | 0.040* |
| Overall management ability | 8.5 (± 0.4) | 7.4 (± 0.6) | 0.76 | 0.019* |

\* Significant result ( $p \leq 0.05$ ). Scores were completed separately for each product; 1 = least favorable rating, 5 = midpoint, 10 = most favorable rating for the product assessed. Values are mean ± SEM. SEM, standard error of the mean; SoC, standard of care.

### Direct Product Comparison

Direct comparison findings showed that 64–82% of participants preferred FLO for hypoglycemia reversal, headache relief, and speed of action (Fig. 1).

**Figure 1.**
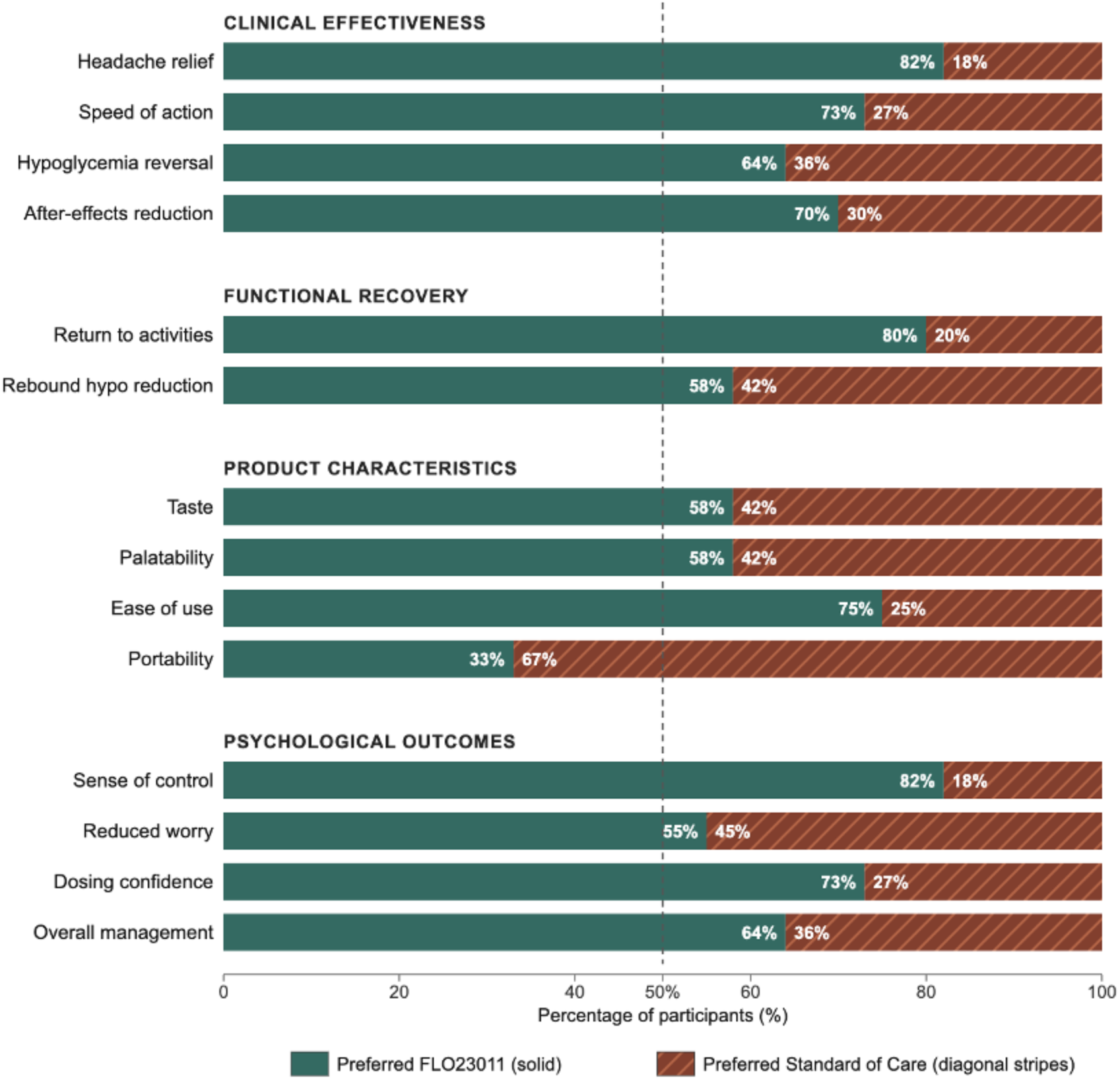
Direct treatment preference. Percentage of participants preferring FLO23011 or standard of care across 14 domains (score > 5 = FLO23011 preference). Preferred FLO23011, solid; Preferred standard of care, diagonal stripes.

Among participants included in recovery-related analyses, 70% reported less severe after-effects such as fatigue or weakness, and 80% reported faster return to daily activities. Rebound hypoglycemia management favored FLO in 58% of participants.

Ease of use was preferred by 75%, while portability favored SoC (33% preferring FLO). Psychological benefits were also reported, including increased sense of control (82%), greater dosing confidence (73%), and reduced worry (55%). Overall, 64% preferred FLO for hypoglycemia management.

Two participants with impaired hypoglycemia awareness and absence of post-hypoglycemic symptoms were excluded from analyses of functional recovery and symptom experience, as these PROs depend on the ability to perceive hypoglycemia and subsequent recovery.

### Qualitative Findings

Closed-response exit interview items showed that 92% of participants rated FLO “Very/Extremely” effective versus 50% for SoC. Similarly, 92% reported “never” or “rarely” experiencing post-hypoglycemia ‘hangover’ symptoms with FLO versus 33% for SoC. More than half of participants described rapid cognitive restoration with FLO, while 75% reported perceived reductions in glucose spikes, which they attributed to more precise dosing and reduced over-treatment.

Open-ended responses were reviewed using an inductive thematic approach. Three themes were identified and are summarized below, with illustrative quotation excerpts provided in Table 2:

- ● Cognitive recovery and mental clarity:

**Table 2.** Structured Exit Interview Themes and Illustrative Quotation Excerpts.

| Theme | Illustrative quotation excerpts |
| --- | --- |
| Cognitive recovery and mental clarity | "Felt more 'with it', more conscious, clearer in my thoughts than normal" [P02]<br>"More switched on and more able to make rational decisions." [P03]<br>"I did not experience the same hangover effect that I usually get following a hypo. Less fatigued and wiped out for the day, and more able to do the things I need to do" [P05] |
| Reduced disruption to daily routine | "The resolution of brain fog did have impact for daily life - just means that you are on the ball and set to go" [P10]<br>"I could return to work quicker." [P04]<br>"It had impact on daily life, took less time out of the day, I could return to normal quicker" [P02] |
| Confidence and control | "Just that confidence: if it is by my bedside or nearby, I just knew it would do the trick," [P04]<br>"It did give me some sort of sense of relief that I knew they were there and they were going to sort me out," [P12]<br>"More confident, more in control." [P07] |

Quotation excerpts are representative verbatim responses from structured exit interviews. Bracketed codes denote anonymized participant identifiers assigned for
*reporting purposes; repeated codes indicate excerpts from the same participant. P, participant.*

Participants described FLO as being associated with faster and clearer mental recovery after hypoglycemia. This theme captured perceived improvements in focus, awareness, mental sharpness, and decision-making capacity. Participants contrasted this with SoC, which was described as being associated with more prolonged cognitive after-effects, including residual “brain fog” and “disorientation.”

- ● Reduced disruption to daily routine:

Participants described FLO as reducing the functional disruption caused by hypoglycemia. This theme reflected faster return to work, social activities, and usual daily tasks, with less prolonged fatigue and less time lost after an episode. By contrast, SoC was described by some participants as leaving them feeling “written off for the day.”

- ● Confidence and control:

Participants described greater confidence in self-managing hypoglycemia with FLO because they perceived it as reliable, effective, and easy to use during an episode. This theme reflected reassurance that the product would resolve the hypoglycemia episode without requiring repeated treatment, excessive decision-making, or major disruption to ongoing activities. SoC was described by some participants as less predictable or reliable.

### CGM-PRO Correlation Analyses

Among participants with both usable CGM data and corresponding questionnaire data (n = 5), reductions in Level 1 hypoglycemia exposure (3.0–3.8 mmol/L) were associated with higher ratings for “reduced worry about hypoglycemia” and “overall hypoglycemia management ability” (Spearman ρ = 0.90; P = 0.037). This association was not observed with SoC and changes in time in the low-normal glucose band (3.9–5.0 mmol/L) were not associated with PRO scores.

Among participants with adequate paired CGM data after quality control (n = 9), baseline glycemic variability predicted the magnitude of CGM-derived response to FLO. Participants with higher coefficient of variation during the SoC period experienced greater improvement in TIR with FLO versus SoC (Spearman ρ = 0.917, P = 0.0005).

### Conclusions

In this randomized, open-label crossover study in adults with type 1 diabetes, FLO was associated with a more favorable patient-reported experience of hypoglycemia recovery than SoC glucose gel. Participants rated FLO more favorably across the majority of assessed domains, including speed of action, reduction in after-effects, return to daily activities, ease of use during hypoglycemia, dosing confidence and overall management ability. Structured exit interviews were consistent with these questionnaire findings, with participants describing faster perceived cognitive recovery, less disruption to daily activities, and greater confidence in self-management with FLO.

The pattern of PRO benefit is consistent with broader evidence that hypoglycemia imposes a burden that extends beyond acute glucose correction. Experimental studies in type 1 diabetes have shown that cognitive recovery after hypoglycemia may lag behind biochemical recovery, with some cognitive domains remaining impaired after plasma glucose normalization (4–6). Evidence also indicates that recurrent hypoglycemia is associated with fear of hypoglycemia and reduced quality of life (8,21,22). The patient-reported domains in which FLO demonstrated benefit therefore correspond to recognized burdens of hypoglycemia. By reporting the full domain-level PRO dataset, structured exit interview themes, and CGM analyses, the present study extends prior glycemic findings, in which FLO significantly improved post-hypoglycemia TIR and reduced recurrent hypoglycemia within 2 hours by 27%, by characterizing hypoglycemia recovery from the patient perspective (20). The alignment of objective CGM data with patient reports of fewer perceived rebounds, greater dosing confidence, and perceived reductions in overtreatment supports the interpretation that FLO may improve the recovery experience after hypoglycemia in ways that are both measurable and meaningful to patients. However, expectancy effects cannot be excluded in an open-label design.

These findings also highlight the behavioral dimension of hypoglycemia recovery. In practice, hypoglycemia is often overtreated (23). This may partly reflect the neurocognitive burden of hypoglycemia, as impaired judgement and delayed cognitive recovery after glucose restoration (4, 24) may make it harder to stop consuming carbohydrate once blood glucose is already recovering. By improving perceived cognitive recovery, reducing after-effects, and increasing confidence that the episode has been adequately treated, FLO may help reduce symptom-driven overtreatment. This may be especially relevant as hybrid closed-loop systems become more widely used, since insulin delivery may already have been reduced or suspended before or during hypoglycemia, altering carbohydrate requirements and increasing the potential for overtreatment to contribute to rebound hyperglycemia (25). The relevance of these findings to reduced-carbohydrate hypoglycemia treatment strategies in hybrid closed-loop users requires further evaluation.

The CGM-PRO findings suggest that psychological benefit may be specifically linked to reduced Level 1 hypoglycemia exposure. Recurrent or symptomatic hypoglycemia is associated with fear of hypoglycemia and compensatory self-management behaviors, including overcorrection, maintaining glucose levels higher to avoid future episodes, and restriction or avoidance of physical activity (26,27). In the present analysis, reduced exposure to Level 1 hypoglycemia was associated with improved psychological PRO scores, whereas the broader low-normal glucose band was not. This pattern suggests that the psychological benefits reported with FLO may reflect reduced exposure to clinically meaningful low-glucose episodes rather than a nonspecific effect of broader glucose changes.

The observed association between baseline glycemic variability and treatment response magnitude suggests that individuals with more unstable glycemic profiles may derive greater objective benefit from MESH treatment. Given the preliminary, hypothesis-generating nature of these findings, they require prospective validation in larger studies. If confirmed, baseline CGM characteristics could help identify individuals most likely to benefit from treatments designed to improve post-hypoglycemia stability and recovery.

A plausible mechanistic explanation for these effects is that BHB provides an alternative neuronal energy substrate during the post-hypoglycemia recovery window. While glucose rapidly corrects plasma glucose levels, neuronal utilization may remain transiently constrained due to NAD⁺ depletion and impaired glycolytic flux (16). BHB is transported across the blood-brain barrier via monocarboxylate transporters, which may be upregulated in diabetes and hypoglycemia (28), and is metabolized downstream of glycolysis. It may therefore bypass this constraint and support ATP production (16,19). Participants’ reports of faster and clearer cognitive recovery are consistent with this hypothesis, although the present study was not designed to test it directly.

Limitations include the open-label design (unavoidable given the organoleptic differences between products) and small sample size, especially for variability-PRO correlation analyses. These analyses were not powered for definitive inference and were not corrected for multiple comparisons. The predominantly male (67%) sample of CGM users may not represent the full type 1 diabetes population. Participants with impaired hypoglycemia awareness or absence of post-hypoglycemic symptoms were excluded from symptom- and recovery-related analyses; therefore, these findings are most directly applicable to individuals who perceive hypoglycemia and post-treatment recovery symptoms.

In conclusion, FLO23011 consistently improved the patient experience of hypoglycemia management across clinical, functional, product-related, and psychological domains, with large effect sizes for ease-of-use and after-effects reduction. These findings complement previously reported glycemic outcomes from the same study and suggest that effective hypoglycemia treatment may extend beyond plasma glucose restoration to encompass functional and experiential recovery. The novel CGM findings suggest that individuals with greater baseline glycemic instability may derive more objective benefit from MESH treatments, and that reduced exposure to symptomatic hypoglycemia may contribute to the psychological gains reported with FLO. Collectively, these observations suggest that binary frameworks for evaluating hypoglycemia treatment based on time-in-range and time-below-range thresholds alone may not fully capture outcomes most relevant to patients. These findings support a broader approach to hypoglycemia self-care, with new thinking needed in three areas: evidence base for treatment products, standards for clinical practice and CGM targets for outcomes.

## Data Availability

The datasets generated during and/or analyzed in the current study are available from the corresponding author upon reasonable request

## Acknowledgements

The authors thank all study participants and acknowledge the support of the Terry Miller Charitable Foundation.

During the course of preparing this work, the authors used KOSMOS (Edison Scientific, edisonscientific.com) for the purpose of supporting focused CGM analyses examining associations between CGM-derived metrics and patient-reported recovery outcomes. Following the use of this tool/service, the authors formally reviewed the content for its accuracy and edited it as necessary. The authors take full responsibility for all the content of this publication.

